# The case of alternatives to opioids: How much do physician characteristics matter when treating a diverse population?

**DOI:** 10.1101/2024.10.15.24315555

**Authors:** Maria L. Alva, Siying Liu

## Abstract

**Rationale:** There is little understanding of the prescription patterns of alternatives to opioids (ALTOS). Monitoring gender and racial health disparities can help with healthcare planning, workforce training, patient education, and awareness.

**Objective:** This study asks whether healthcare professionals, when treating patients of the same race and sex, increase their likelihood of prescribing ALTOs relative to opioids.

**Methods:** We use national Medicare Part D data from 2013 to 2017 and a machine-learning algorithm informed by census data to define the race of prescribers. We use multivariate regression models to understand the impact of race and sex biases on the extensive margin (e.g., percentage of people receiving ALTOs) and the intensive margin (e.g., the number of ALTOs prescriptions per capita).

**Results:** Between 2013 and 2017, there has been an 8.7% increase in the prescriptions of ALTOs. The number of beneficiaries receiving ALTOs increased by 11.4%. In 2017, the number of ALTOs prescriptions per capita written as a fraction of all painkillers was 45%, and the number of beneficiaries receiving ALTOs prescriptions as a fraction of people receiving at least one form of painkillers (ALTOs or opioids) was 49%. A male doctor is 20.4% more likely to prescribe ALTOs as the percentage of same-sex patients increases. A white doctor is 7.4% more likely to prescribe ALTOs as the percentage of same-race patients increases, even when controlling for the socioeconomic status of patients, their age and risk factor, and the state and specialty of the prescriber.

**Conclusion:** Sex and race concordance between providers and patients are significantly associated with prescribing alternatives to opioids. These systematic differences could be addressed by supporting diversity in the workforce, training, and increasing the minimum amount of time a visit should last.

## Introduction

Chronic pain is defined as pain that lasts more than three months or past the time of normal tissue healing, and it can affect a person’s quality of life significantly. Prescriptions of opioids such as oxycodone, morphine, and codeine help treat painful conditions but can increase the risk of addiction, overdose, and death when taken for prolonged periods and at high doses. There is now consensus that for some painful conditions, opioids should not be the first line of treatment For example, chronic conditions such as headaches and back pain can be made worse when opioids are used because people develop tolerance to opioids requiring increasingly higher dosages to achieve the same effect. The CDC considers opioids to be inadequate first-line treatments for chronic pain and that providers are the best line of defense for preventing opioid overuse and addiction when alternatives exist (2).

Alternatives to opioids (ALTOs) are regarded as safe substitutes for alleviating pain and a tool that healthcare providers can use to curb opioid over-prescription. ALTOs can be pharmacological (e.g., topical analgesics, muscle relaxants, anticonvulsants, and nonsteroidal anti-inflammatory drugs [NSAIDs]) or non-pharmacological options (e.g., physical therapy, acupuncture, and other relaxation techniques) (3). Pilot programs such as the Alternatives to Opioids (ALTOs) program in New Jersey (4) and Colorado (Colorado Hospital Association) have shown that with clear provider guidelines and protocols, prescribing ALTOs is effective in managing pain and reducing opioids use and dependency in surgical and Emergency Department settings.

Monitoring gender and racial health disparities and biases is important for healthcare planning, workforce training, patient education, and awareness. Little research has been done on classifying and understanding the prescription patterns of ALTOS (2). Changes in the demographic distribution of opioid-related morbidity and mortality warrant greater attention to prescription practices across a diverse population as the US. The contributions of this study are threefold: (1) oo define a list of drugs prescribed as alternatives to opioids. (2) to understand the likelihood of receiving alternatives to opioids based on both race and sex matching between providers and patients; and (3) to understand the impact of race and sex biases on both the extensive margin (e.g., percentage of people receiving ALTOS as a fraction of all painkillers) and the intensive margin (e.g., how many prescriptions of ALTOS per capita, on average, are written).

### Evidence of the changing demographics of opioid-involved overdose death rates

Case and Deaton (2015) (5) showed that unparalleled access to prescription opioids caused a decline in life expectancy for White people. The decrease in White life expectancy began in 1998, two years after the US Food and Drug Administration approved OxyContin. At its highest, Healthcare providers wrote over 250 million painkiller prescriptions yearly (6). The promotion, marketing, and distribution of opioids had significant geographical variations. Alabama, Kentucky, Maine, Virginia, and West Virginia saw up to five times more prescriptions than the national average (7). A 2012 meta-analysis of the evidence on racial/ethnic disparities in pain treatment in the United States showed that Blacks/African Americans were under-treated (8).

This type of evidence led some commentators to conclude that Blacks/African Americans “benefited” from lack of access to pain treatment and that, in the case of opioids, racial bias might have inadvertently “protected” this group (9). Recent research has focused on the convergence of opioid misuse in Black and White populations. Between 2013 and 2015, mortality due to opioids increased at an annual rate of 18% (95% CI = 1%, 39%) for White and 34% (95% CI = 2%, 77%) for Black people (10). There are many facets to the opioid epidemic that might have contributed to the convergence between Whites and Blacks; from the use of heroin as a substitute to pills when prescription drugs became too expensive; to the rise in the use of fentanyl, a synthetic opioid available both as an illegal street drug and as a prescription drug to treat patients with severe pain or resistant to weaker opioids (11). White patients are significantly more likely to receive prescriptions for medications to help with withdrawal symptoms (e.g., buprenorphine and naloxone) compared to African Americans (12). Collecting data on and monitoring recurring differences in treatment as new protocols in the pharmacological management of chronic pain become available, including ALTOs, becomes essential to understand whether standards of care adhere to protocols or are unequal.

### Evidence on how the pairing of doctors and patients by race and sex affects outcomes

Previous research has shown that disparities in health outcomes can be linked to social determinants and provider bias. For example, doctor-patient commonalities seem to affect both health and behavioral outcomes. There is increasing evidence that female patients receive better care from female physicians than male physicians. Female patients randomly assigned to a female doctor were 8.5% more likely to be evaluated as having a disability and receive compensation after an injury than when evaluated by a male doctor (male and female doctors evaluated male patients consistently) (13). Female patients treated by male physicians have a small but statistically significant increased likelihood of adverse postoperative outcomes (deaths, readmission, and complications within 30-day following surgery) compared to males treated by male or female surgeons (14). Gender concordance increases a patient’s probability of survival after a heart attack by 5.4%. Mortality rates decrease when male physicians practice with more female colleagues or have previously treated more female patients (15).

The race concordance between patients and doctors also appears to matter for health outcomes. When Black newborns are cared for by Black physicians, the mortality penalty they suffer, as compared with White infants, is halved (16). When Black men are randomized to Black instead of non-Black male doctors, they are more likely to select preventive services (17). A recent study found that 8.2% of patients in a large academic and metropolitan medical center had negative descriptors in their medical notes and that Black patients had 2.5 times the odds of having such descriptors. (18) Patients on Medicaid or Medicare also had higher odds of negative descriptors than those with commercial insurance, and the same was true for unmarried patients compared to married ones. The likelihood of negative descriptors decreases in outpatient notes compared with emergency department and hospitalization notes, suggesting that getting to know patients better decreases biases. 17% of Medicare patients feel vulnerable to prejudice in the healthcare setting due to their race or gender. Patients who feel vulnerable are worse off health-wise and less likely to seek treatment. (19)

Race and gender concordance matter because of bias and prejudice, whether these are voluntary or not. For example, doctors might involuntarily stereotype patients (based on race or gender) when making quick decisions. Research has shown that even unprejudiced physicians rely on prior probabilities more often than current information when treating patients of a race different than their own, possibly due to miscommunication and misunderstanding resulting from cultural differences. (20)

## Methods

We study the association between physician characteristics (race, sex) and beneficiary characteristics (race, sex, age, dual status, and health risk score) with the ALTOs prescription rate, controlling for rural/urban location and disease specialty. We explore whether Medicare prescribers are more likely to prescribe ALTOs relative to opioids to a patient who is of the same race or sex as themselves. Or if race and sex are “catch-all” variables for other factors such as income and education (via dual status, i.e., the patient is both Medicare and Medicaid eligible) and other behavioral factors (via risk scores).

### Data

We use Medicare Part D Prescriber Public Use File (PUF) (21) data from 2013 to 2017. PUF is a prescriber-level data set containing Prescription Drug Event records submitted by Medicare Advantage Prescription Drug plans and stand-alone Prescription Drug Plans. We identify individual providers and their specific prescriptions. We compare the prescription of opioids and pharmacological ALTOs. The list of ALTOs and opioids is available in Appendix A.

We complement the PUF with two other data sources: (1) the Rural-Urban Commuting Areas taxonomy (22) using a ZIP code crosswalk. We expect rural populations to be more vulnerable to opioids than their urban counterparts because of longer travel distances to— and higher costs associated with— health care services and a greater emphasis on generalists. (2) Census data.

Claims data capture the race of beneficiaries but not the race of providers. To define the race of prescribers, we used a machine-learning algorithm based on prescribers’ first and last name and trained on census data (Python package “ethnicolr”) (23).

### Measures and Analysis

We compute two outcomes (*y*_*p,t*_) of interest, defined at the level of the provider:

I. # ALTOSs claims / (# ALTOSs claims + # Opioid claims)

II. # ALTOSs beneficiaries / (# ALTOSs beneficiaries + # Opioid beneficiaries)

where # means number. Our model is specified as follows:

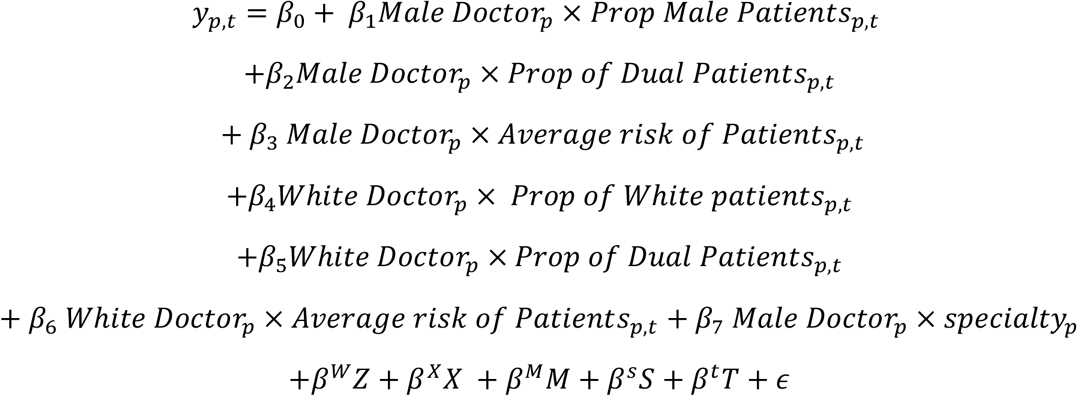

Where p denotes each prescriber and t denotes the year. The vector **Z** includes the following prescriber characteristics: sex, race, location, and specialty. The vector **X** represents the characteristics of the prescriber’s patients (proportion of male patients, race, younger than 65 and older than 65, dual-eligible, and average health risk score). **M** denotes the medical specialty of the provider. We include the male doctor by specialty interaction to account for the selection in sex across specialties. **S** denotes the state-fixed effects and **T** denotes year dummies.

We test two versions of the model, with and without the interaction of the dual beneficiary status and the patient risk score and with the doctor’s sex (*β*_2_ and *β*_3_) and race (*β*_5_ and *β*_6_). These additional controls are essential because the risk score indicates the patient’s health, and dual status is a proxy for the patient’s socioeconomic status. Risk scores represent the deviation in average spending of patients compared to the age and sex-adjusted overall index for the entire Medicare population. We, therefore, compare the full model with a parsimonious model that does not control for the interactions of the patient risk score with the doctors’ race and sex and the interactions of the dual status with the same variables. The parsimonious model might capture correlations on race and sex that under the full model might be explained by the risk profile and the socio-economic status of patients.

*β*_1,_ and *β*_4_ are the key parameters of interest, which can be interpreted as the likelihood of treating patients of the same sex and the same race with ALTOs compared to opioids.

## Results

Exhibit 1 shows the summary statistics for the 2013-2017 empirical sample representing 415,464 Medicare prescribers nationally. The sample comprises the subset of providers that prescribed both ALTOs and opioids – 40% of patients served during this period were male, 77% Caucasians, and 32% duals. The average risk score among Medicare beneficiaries prescribed opioids or ALTOs was 1.42. As a point of reference, a risk score of 1 reflects an average patient, and therefore 1.42 reflects a patient population with a cost of care 42% higher than the average. Thirty nine percent of Medicare beneficiaries were individuals older than 75. There is a non-negligible fraction of individuals younger than 65 (23%); this subset of the sample represents individuals with disabilities.

**Exhibit 1:** Provider and Beneficiary Characteristics, 2013-2017

| Variable | Mean | Std.<br>Dev. |
| --- | --- | --- |
| **Medicare Patient characteristics** |  |  |
| % of male patients | 39.8% | 10% |
| % White patients | 77.4% | 24% |
| % Hispanic patients | 5.3% | 15% |
| % Black patients | 6.8% | 16% |
| % Asian patients | 1.3% | 8% |
| % of duals served | 31.5% | 23% |
| Average risk* | 1.42 | 0.55 |
| % patients <65 | 23.4% | 19% |
| % patients >75 | 39.4% | 11% |
| **Doctor characteristics** |  |  |
| % White doctors | 79.6% | 40% |
| % Asian doctors | 13.0% | 34% |
| % male doctors | 65.9% | 47% |
| urban | 89.4% | 31% |
| **Outcomes (2013)** |  |  |
| # ALTOSs claims / (# ALTOSs claims + # Opioid claims) | 33.7% | 36.2% |
| # ALTOSs bene / (# ALTOSs bene + # Opioid bene) | 40.4% | 34.4% |
| **Outcomes (2017)** |  |  |
| # ALTOSs claims / (# ALTOSs claims + # Opioid claims) | 45.1% | 39.2% |
| # ALTOSs bene / (# ALTOSs bene + # Opioid bene) | 49.1% | 36.8% |

\* Average hierarchical condition category risk score of beneficiaries, as computed by CMS for provider reimbursement.

Roughly 80% of doctors had Caucasian surnames, based on the census matching algorithm we used. The algorithm aggregates names and surnames into six possible racial and ethnic groups. 13% of doctors were Asian. The remaining 7.4% of doctors were Black, Hispanic, American Indian, Alaska Native, Native Hawaiian, and Other Pacific Islander or were classified as having two or more races. 89% of doctors operated in an urban setting.

Between 2013 and 2017, there has been an increase in 8.7 percentage points in the prescriptions of alternatives relative to opioids in Medicare beneficiaries. The number of beneficiaries receiving an ALTO prescription as a fraction of people prescribed both ALTOs and opioids have also increased, at a higher rate, 11.4%. In 2017, the number of ALTOS prescriptions per capita written as a fraction of all painkillers was 45%, and the number of beneficiaries receiving ALTOS prescriptions as a fraction of people receiving at least one form of painkillers (ALTOS or opioids) was 49%.

Exhibit 2 shows the regression results of the full model (model 2) and the parsimonious model (model 1) for the two outcomes of interest. The parsimonious model reports larger coefficients on the parameters of interest (*β*_1,_ and *β*_4_, model 1, columns 2 and 4) compared to model 2 (columns 3 and 5). We include but do not show in the table state, year, and specialty fixed effects. For both outcomes (i.e., intensive and extensive margin), ALTOs prescription ratios are higher if the prescriber has a higher percentage of patients of the same sex and race, with sex being a greater differentiator of treatment options than race, in the parsimonious model.

**Exhibit 2:**
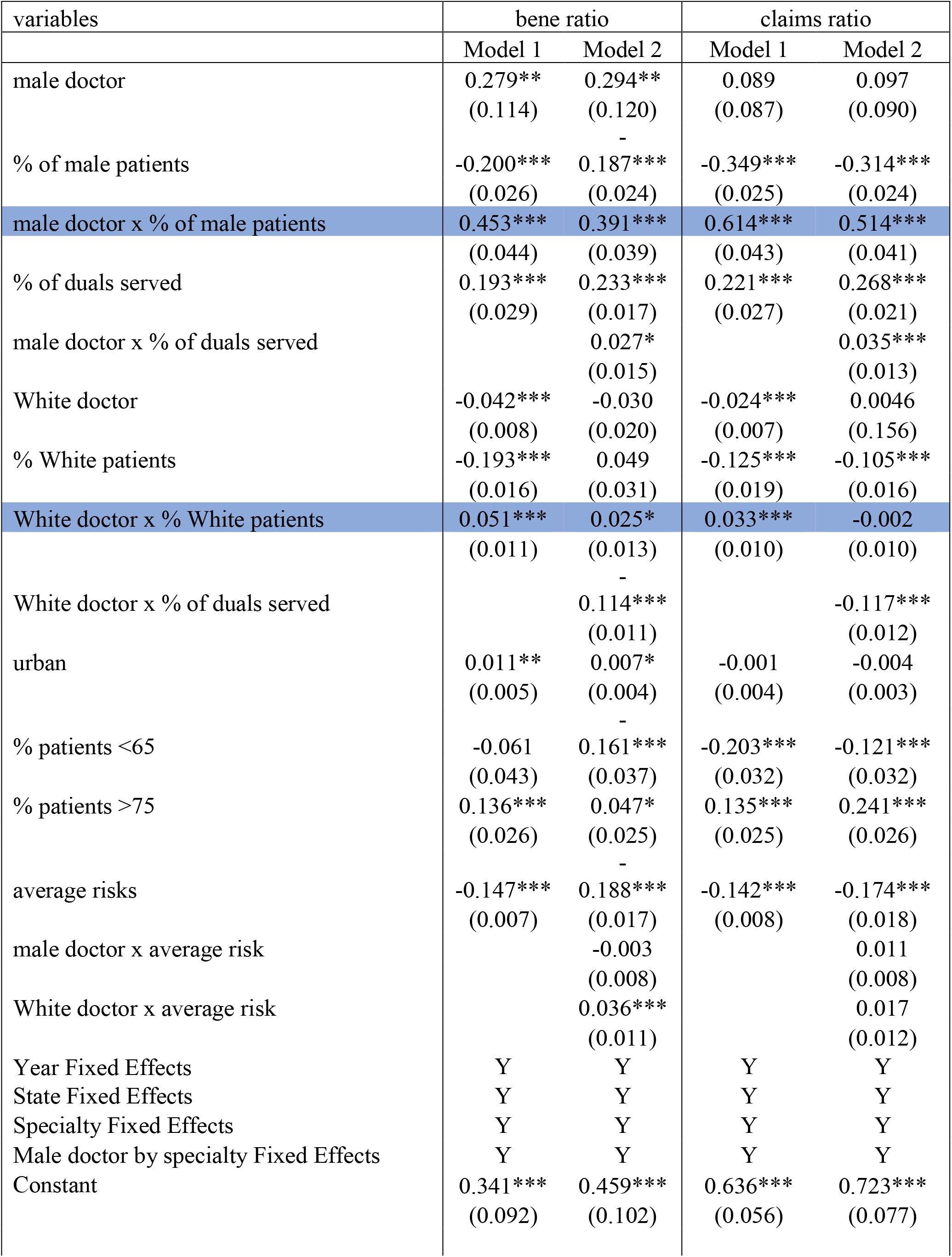
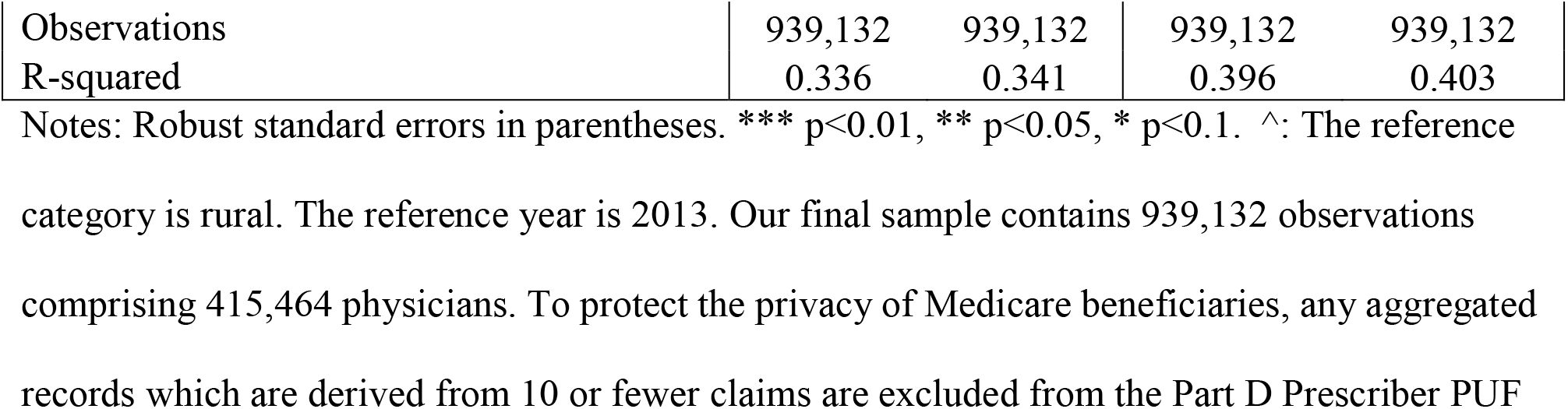
Parsimonious (model 1) and full (model 2) multivariate regression models

A male doctor is more likely to prescribe ALTOS than a female doctor. A female doctor would prescribe 18.7% less ALTOS relative to all forms of painkillers to male patients. A male doctor is 20.4% more likely to prescribe ALTOs to a male patient (39.1%-18.7%). A male doctor would also increase the quantity of ALTOs prescribed to male patients proportionately by 20% (51.4-31.4). A female doctor would decrease the number of ALTOs relative to all other forms of painkillers by 31.4%. Both coefficients related to the number of people and the number of claims are statistically significant at the 0.01 level.

A White doctor is less likely to prescribe ALTOs than a doctor of another race or ethnicity. Race matters more in the parsimonious model (model 1) than in the model that controls for the interaction between the doctor’s race with the patients’ dual status and average risk score (model 2). White doctors are less likely to prescribe ALTOS to duals and patients with high-risk profiles than non-White doctors. However, a white doctor is 7.4% more likely to prescribe ALTOs to a white patient (2.5%+ 4.9%) than a patient of a different race. A white doctor is less likely to increase the quantity of ALTOs prescribed to white patients. The intensive margin shows a 10.7% (−10.5%-0.2%) decrement in the number of ALTOS relative to other pain relief options.

Across all outcomes, ALTOs ratios are higher if the prescriber has a higher proportion of dual-eligible beneficiaries, older than 75 and having a lower health-risk score. ALTOs’ prescriptions disproportionately concentrate on certain specialties such as endocrinology (Appendix B).

## Discussion

Disparities in pain management based on prescriber and patient sex and race should not exist nor determine the quality of care a patient receives. Our findings show that health care providers’ sex and race are significantly associated with prescribing alternatives to opioids. While further work should focus on understanding the reasons for these differences, differences in needs and access may exist across sex and race that may be correlated with differences in treatment. This paper controls, to some extent, for socio-economic factors via the indicator variable for duals. Patients’ health risks and dual status confound racial discrepancies in care. Dual status accounts, to some extent, for race but not sex disparities. Anecdotal evidence from physicians working in the ER corroborates our findings regarding dual beneficiaries. Doctors are more cautious about prescribing opioids to individuals they think are likely to resell them illicitly. It is interesting, however, that in our data, White doctors appear to be more affected by the dual status of patients than non-White doctors. We interpret this finding as evidence of structural and institutional racism rather than internalized or interpersonal racism.

We find more significant discrepancies in the treatment of males and females than across races. Male doctors might have more difficulty interpreting the clinical manifestations of pain in female patients than male ones, possibly resulting from stereotypes related to the so-called “weaker sex”(24). Sex-driven differences in care persist regardless of these health and socio-economic proxies.

Health care service and outcome data can help address health equity. While outward similarities between doctors and patients may increase the level of trust and communication achievable in clinical practice, systematic differences in health have the power to produce health inequity. The presence of sex and racial disparities in prescriptions suggests at least three avenues for action:

(1) Healthcare systems must invest in supporting diversity in the workforce. Given that female and non-White doctors are underrepresented in the physician workforce, we should invest in supporting the next generation of physicians to serve the communities that they represent. (2) Training efforts to combat prejudice and eliminate stereotypes could help, as well as training on protocols regarding criteria for treatment by condition. (3) The care system should be designed so that healthcare visits allow a minimum adequate amount of time for good communication to emerge. Increasing the minimum amount of time a visit should last could help address some communication barriers between doctors and patients.

### Limitations

There are four possible limitations to our findings and their interpretation. All four limitations are data-driven. First, while part D Prescriber PUF has a wealth of information, it may not represent all Medicare patients. PUF only includes information on beneficiaries enrolled in Medicare Part D (i.e., approximately two-thirds of all Medicare beneficiaries) (25).

Second, the fact that we do not observe the race/ethnicity of the prescriber might add noise to the estimated associations. The accuracy of the predicted race/ethnicity depends on the power and reliability of the census-trained algorithm. The algorithm we used has an out-of-sample precision and recall of .83 and .84, respectively (23).

Third, the PUF is a provider-level data set. The fact that we do not observe patient-level outcomes might give rise to an ecological fallacy, which might impact the interpretation of results. For example, the results assume that the only drivers in interpreting *β*_1_ and *β*_4_ coefficients are the doctors’ sex and race rather than the composition of the population mix. For example, a White doctor serving 50% White patients and a White doctor serving 90% White patients might not necessarily be 1.25% (2.5%*50%) and 2.25% (2.5%*90%) more likely to prescribe ALTOs to a White patient, compared to a non-White doctor because a positive correlation at the aggregate level does not necessarily translate to a positive correlation at the individual level.

Finally, a more comprehensive understanding of the perceived differences across sexes in pain sensitivity and the different levels of exposure, recovery, and use of alternatives is needed.

## Data Availability

All data produced in the present work are contained in the manuscript.

## Appendix A: Generic Drug Names used to define the ALTOs

**The list of generic ALTOs used in this analysis are**: acetaminophen, amitriptyline, anaprox, bupropion, carbamazepine, celecoxib, citalopram, cyclobenzaprine, dexamethasone, diazepam, diclofenac, diflunisal, diprivan, duloxetine, garbapentin, etodolac, fenoprofen, flurbiprofen, haloperidol, ibuprofen, ketamine, ketoprofen, ketoroloc, lidocaine, meclofenamate, meloxicam, metoclopramide, nabumetone, naproxen, oxaprozin, piroxicam, sulindac, sumatriptan, tizanidine, tolmetin, and venlafaxine.

Note: some drugs like propofol and ropivacaine are not covered under Medicare part D and are therefore not part of the ALTO’s definition in Medicare.

The Centers for Medicare & Medicaid Services release in the Prescriber Public Use file (PUF) an indicator variable that flags the number and type of opioid prescriptions administered. The list of opioids used in this study is the one reported in PUF and available here: https://www.cms.gov/research-statistics-data-and-systems/statistics-trends-and-reports/medicare-provider-charge-data/part-d-prescriber.html

## Appendix B: ALTOS/ Opiods Claims by specialty

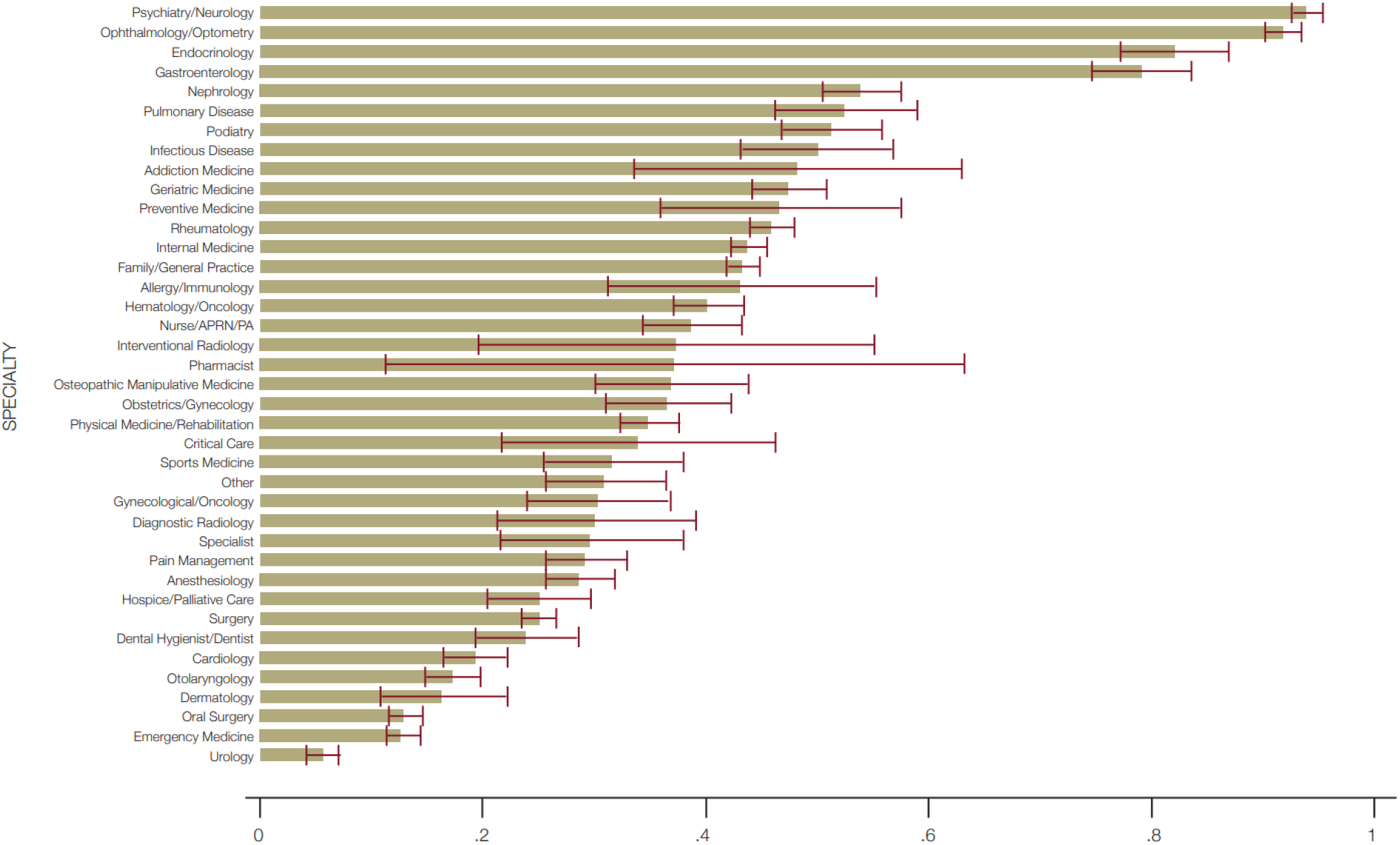

